# Contributions of Genetic Liability and the COVID-19 Pandemic to Rising Psychopathology Among Youth in the United States

**DOI:** 10.1101/2025.04.14.25325727

**Authors:** Lucy Shao, Jonathan Ahern, Robert Loughnan, Bohan Xu, Holly E. Baker, Susan F. Tapert, Fiona C. Baker, Wesley K. Thompson, Orsolya Kiss, Eva M. Müller-Oehring, Marie Gombert-Labedens, Chun Chieh Fan

## Abstract

**Background:** Adolescent mental health issues were surging during COVID-19 pandemic. Yet it is unclear whether the pandemic amplified pre-existing vulnerabilities for psychiatric disorders.

**Methods:** Using the longitudinal data from Adolescent Brain Cognitive Development (ABCD) Study^©^ (n = 7,560, 2∼3 waves of assessments before pandemic and 2∼3 waves after first nation-wide pandemic lock-down), we evaluated associations of the pandemic, genetic liabilities to psychiatric disorders, and their interactions with 20 different measures of psychopathology. Genomic common factor models aggregated genomic effects across eight psychiatric disorders, summarizing into four latent factors. Analyses were stratified by genetic ancestry and sex.

**Results:** In European-like ancestry adolescents, each 1 standard deviation increase in Neurodevelopmental (ND) or Internalizing (INT) PRS significantly associated with increment in most psychopathologies by 3% to 19%. After controlling for individuals PRS, pandemic periods were significantly associated with accelerated rates in parent-reported Child Behavior Checklist (CBCL) withdrawn depressed and rule-breaking syndrome scores, CBCL DSM-oriented conduct, somatic, and attention deficit/hyperactivity problems, and all corresponding youth-reported Brief Problem Monitor (BPM-Y) scores. In sex stratified analysis, CBCL DSM-oriented affective problem scores significantly worsened in early pandemic among females (21% increase; 95%CI 13%-29%; P=2.6×10□□) but not males. Females have stronger associations between INT PRS and rate of increment on CBCL DSM-oriented affective problem scores (15%; 95%CI 10%-21%; P=6.4×10□¹□), comparing to males (10%; 95%CI 6%-15%; P=4.2×10□□). The multiplicative interactions between PRS and pandemic periods were at-most trending, showing positive interactions between ND PRS and early pandemic among females for CBCL conduct (15%; 95%CI 7%-23%; P=5.17×10□□) and aggressive behavior scores (9%; 95%CI 4%-14%; P=5.09×10□□).

**Conclusion:** A wide range of adolescent mental health symptoms intensified during the pandemic period. Both genetic vulnerabilities and pandemic-related factors are associated with increased psychiatric symptoms. The genetic liability and the pandemic periods were associated with mental health issues independently, meaning genetically at-risk individuals saw a higher relative increase in mental problems during the pandemic. Females exhibited higher levels of mental health symptoms and more sustained increases across the duration of the pandemic compared to males. Youth with genetic vulnerability to neurodevelopmental phenotypes required special attention due to heightened mental health risks during stressors like COVID-19.

## Introduction

The COVID-19 pandemic brought unprecedented challenges worldwide, significantly impacting mental health across various demographics. Among youth, the risk of having depression in the first few months of pandemic was reported to be three times higher than during the pre-pandemic time (Pelham et al., 2021). Global estimates showed that the prevalence of depression and anxiety among children and adolescents increased by 2-fold, from 11-12 percent to 20-21 percent in the first year of the COVID-19 pandemic (Racine et al., 2021). Social isolation, disrupted routines, academic pressure, fear of infection, and economic strain may all contribute to the deterioration of mental health among young individuals (Guessoum et al., 2020; Loades et al., 2020; Pfefferbaum & North, 2020; Wang et al., 2020).

While evidence of poor mental health outcomes among youth in the first year of COVID-19 is comprehensive, it is unclear if these effects held into the second year of the pandemic, when vaccines began to be rolled-out, lockdown efforts became uneven, infection-incidence waxed and waned, and youths’ routines adapted (Campione-Barr et al., 2024; Marshall et al., 2022; Morales-Vives et al., 2024; Viner et al., 2022). Such adaptations and changing socio-environmental challenges from the pandemic may have resulted in less depressive and anxiety symptoms among youth. Conversely, mental health problems may have lingered, as social disruptions may have had accumulative impacts regardless of the changing pandemic landscape.

Critically, the rise of mental health issues among some youth may have been driven by pre-existing vulnerabilities toward psychiatric disorders (Kiss et al., 2022, 2024). Most of the discussion regarding groups sensitive to the unique challenges of COVID-19 pandemic has focused on socio-economic disadvantages as risk factors (Adise et al., 2024; Jones et al., 2022; Mir M. Ali, Creedon, Disability, & Evaluation., 2022; Pfefferbaum & North, 2020; Pierce et al., 2020). Genetic liability for mental health problems among youth have been largely ignored despite that psychiatric disorders have a considerable heritable component (Adise et al., 2024). How COVID-19 pandemic challenges interacted with genetic pre-dispositions remain unclear. The rising prevalence of the mental health issues may have resulted from independent effects of pandemic related socio-environmental challenges and genetic liabilities, or it may have resulted from disruptions accelerating disease processes among youth at high risk due to genetic and environmental liabilities, resulting in higher incidences for these individuals.

To answer these questions, we utilized longitudinal data from a large-scale adolescent cohort, the Adolescent Brain Cognitive Development^SM^ (ABCD) study. The COVID-19 pandemic happened during the longitudinal follow-up of ABCD Study^©^ when the participants of age range from 10 to 14 at 2020. Given the sample size, interlacing visits with pandemics, and the availability of the genomic data, we can investigate associations and interactions of different mental health challenges faced by adolescents during the COVID-19 pandemic overtime.

## Method

### Study cohort

This study uses ABCD Study data release 5.0. The ABCD Study–the largest in the U.S. assessing longitudinal brain development–was initiated by the United States National Institutes of Health (NIH) in 2015. Starting aged 9-10 years, children are being followed annually with comprehensive measures, including both self- and parent-report questionnaires (Garavan et al., 2018; Volkow et al., 2018). More details about the ABCD Study can be found at http://abcdstudy.org.

From 2016-2023, there were five assessments of general psychopathology: parent reports from the Child Behavior Checklist (CBCL) and self-reports from the Brief Problem Monitor (BPM-Y). Of 11,880 participants, 9,875 had at least the one assessment while 7,560 of them have genetic data available. A visualization of data distribution by time is presented in **Appendix Figure S1**. Given that we use polygenic risk scores (PRSs) throughout our analyses in this report, which are known to have significant challenges of portability between ancestral groups (Choi, Mak, & O’Reilly, 2020), we stratified eligible individuals into two sub-cohorts, each analyzed independently. EUR is our primary study cohort since their genetic background is matched with the study populations of the genome-wide association studies (GWAS) which the PRS are based upon. DIV is our secondary analytic group because the increased inferential uncertainties given mismatched study populations between the GWAS training samples and the PRS application sample (Ding et al., 2023). Table 1 outlines the demographics of the two samples.

**Table 1.** Demographic Characteristics of the Two Sub-Cohorts.

| Characteristic | European-like(EUR)<br>ancestry (n = 4,750) | Non-European-like<br>ancestry (n = 2,756) |
| --- | --- | --- |
| **Baseline Age, mean (SD)** | 9.96 (0.62) | 9.92 (0.62) |
| **Female** | 2,228 (46.9%) | 1,327 (48.1%) |
| **BMI, mean (SD)** | 17.9 (3.36) | 19.8 (4.43) |
| **Parents Married** |  |  |
| └ Yes | 3,900 (82.1%) | 1,436 (52.1%) |
| └ No | 847 (17.8%) | 1,283 (46.6%) |
| **Household Income** |  |  |
| └ < \$50,000 | 584 (12.3%) | 1,216 (44.1%) |
| └ \$50,000–\$99,999 | 1,405 (29.6%) | 672 (24.4%) |
| └ ≥ \$100,000 | 2,539 (53.5%) | 542 (19.7%) |
| **Parent's Highest Education** |  |  |
| └─ < High School Diploma | 231 (4.9%) | 319 (11.6%) |
| └─ High School Diploma/GED | 132 (2.8%) | 338 (12.3%) |
| └─ Some College | 806 (17.0%) | 931 (33.8%) |
| └─ Bachelor's Degree | 658 (13.9%) | 330 (12.0%) |
| └─ Postgraduate Degree | 2,145 (45.2%) | 567 (20.6%) |
| **Self-Reported Race at Baseline** |  |  |
| └─ White | 4,491 (94.5%) | 629 (22.8%) |
| └─ Black | 4 (0.1%) | 925 (33.6%) |
| └─ Asian | 4 (0.1%) | 156 (5.7%) |
| └─ Mixed | 196 (4.1%) | 672 (24.4%) |
| └─ AIAN/NHPI | 13 (0.3%) | 28 (1.0%) |
| └─ Other | 26 (0.5%) | 271 (9.8%) |

### Child Behavior Checklist (CBCL)

The CBCL (Achenbach, 2009) is based on the Achenbach System of Empirically Based Assessment (ASEBA), which measures behavioral and emotional problems among children and adolescents. CBCL was completed by parents/caregivers (Mazefsky, Anderson, Conner, & Minshew, 2011). resulting in eight syndrome (Figure S2) scales (anxious/depressed, withdrawn/depressed or depressed, somatic complaints, social problems, thought problems, attention problems, rule-breaking behavior, and aggressive behavior) and six DSM-oriented (Figure S3) scales (affective problems, anxiety problems, somatic problems, attention deficit/hyperactivity problems, oppositional defiant problems, and conduct problems). CBCL DSM affective problems is specifically designed to assess symptoms related to depressive disorders, so we refer it as DSM depress in this paper. Two additional higher-order summary scores were derived based on syndrome scales and raw scores, i.e. internalizing and externalizing problem scores. We used the raw summary scores of CBCL across these syndrome and diagnostic scales. Therefore, in total, we examined 16 different CBCL scores as the main outcomes in this study.

### Youth Brief Problem Monitor (BPM-Y)

The BPM forms are developed from the longer corresponding versions of the ASEBA *Child Behavior Checklist for those aged* 6–18 *years*. To evaluate the subjective well-being of ABCD Study participants, we also used self-rated Youth Brief Problem Monitor BPM-Y(Achenbach, 2009) questionnaire. It contains four summary measures, based on children’s self-report, including Total Problems, Internalizing Problems, Externalizing Problems, and Attention Problems(Figure S5). In total, we examined 4 different BPM-Y scores as outcomes in this study.

### Classification of early pandemic and late pandemic period

To examine patterns of change over the course of the pandemic, we used January 2021 as the demarcation between “early” and “late” pandemic periods. We defined the early pandemic period as starting from March 2020, when lock-down orders in the U.S. were announced, to December 2020, when the first COVID-19 vaccines appeared on the horizon. The late pandemic period was defined as the entire year of 2021 since several waves of outbreaks were still happening and COVID-19 social distancing measures were executed unevenly across the U.S. The reference is the pre-pandemic state (before March 2020).

### Genotype data and analysis

Genetic information was retrieved from saliva and blood sources and assayed using the Smokescreen™ Genotyping array (Baurley et al., 2016; Uban et al., 2018). Using HRC reference panel with TOPMED imputation server (Taliun et al., 2021), the genotypes were imputed in 280,850,795 imputed variants, aligned to genome build GRCh38. Genetic principal components were calculated using PC-Air (Conomos et al., 2015) with default settings. Using PLINK (v2) (Chang et al., 2015), we filtered the input data to include only autosomal variants with a minor allele frequency of 1% or greater, which left us with 10,927,293 variants.

### Genetic ancestry calculation

Genetic continental ancestry was estimated using SNPweights (Chen et al., 2013) and external genomic reference panels for African, East Asian, European (Auton et al., 2015), and Indigenous North and South American (Reich et al., 2012) ancestry populations. ABCD participants were separated into continental ancestry groups if they had an inferred genetic ancestry of at least 80% consistent with a continental reference panel; if not, they were assessed as having admixed ancestry. Using inferred genetic ancestries allowed us to more precisely define admixture in the ABCD cohort; previous work has shown that ABCD inferred ancestries closely align with genetic principal component analysis (Ahern et al., 2023). The genetically derived ancestral categories were used to define the study subsets, i.e. EUR and DIV.

### Latent factor polygenic scores for psychiatric disorders

To maintain a comprehensive characterization of psychopathological liabilities using genetic information while reducing the burden of multiple testing, we used the framework developed by Grotzinger et al (Grotzinger et al., 2022). Using the structural equation model formulation implemented in Genomic SEM, we fit confirmatory factor models based on summary statistics of GWAS of 8 psychiatric disorders: attention-deficit/hyperactivity disorder (ADHD) (Demontis et al., 2019), anorexia nervosa (AN) (Watson et al., 2019), autism spectrum disorder (AUT) (Grove et al., 2019), anxiety disorders (ANX) (Otowa et al., 2016), bipolar disorder (BIP) (Stahl et al., 2019), major Depressive disorder (MDD) (Wray et al., 2018), obsessive-compulsive disorder (OCD), and schizophrenia (SCZ) (Trubetskoy et al., 2022). The best fitting model (Comparative Fit Index (CFI) of 0.984) resulted in four latent factors: Compulsive disorders (COM), Psychotic disorders (PSY), Neurodevelopmental disorders (ND), and Internalizing disorders (INT). The first factor, COM consists of disorders characterized by compulsive behaviors AN and OCD. The second factor, PSY factor characterizes disorders that may had psychotic features SCZ, BIP, and MDD. The third factor, ND factor represents disorders with developmental origins, i.e. ADHD and AUT. The fourth factor, INT factor characterizes ANX and MDD, which have internalizing symptoms. The four-factor model structure and coefficients are shown in **Figure 1**.

**Figure 1:**
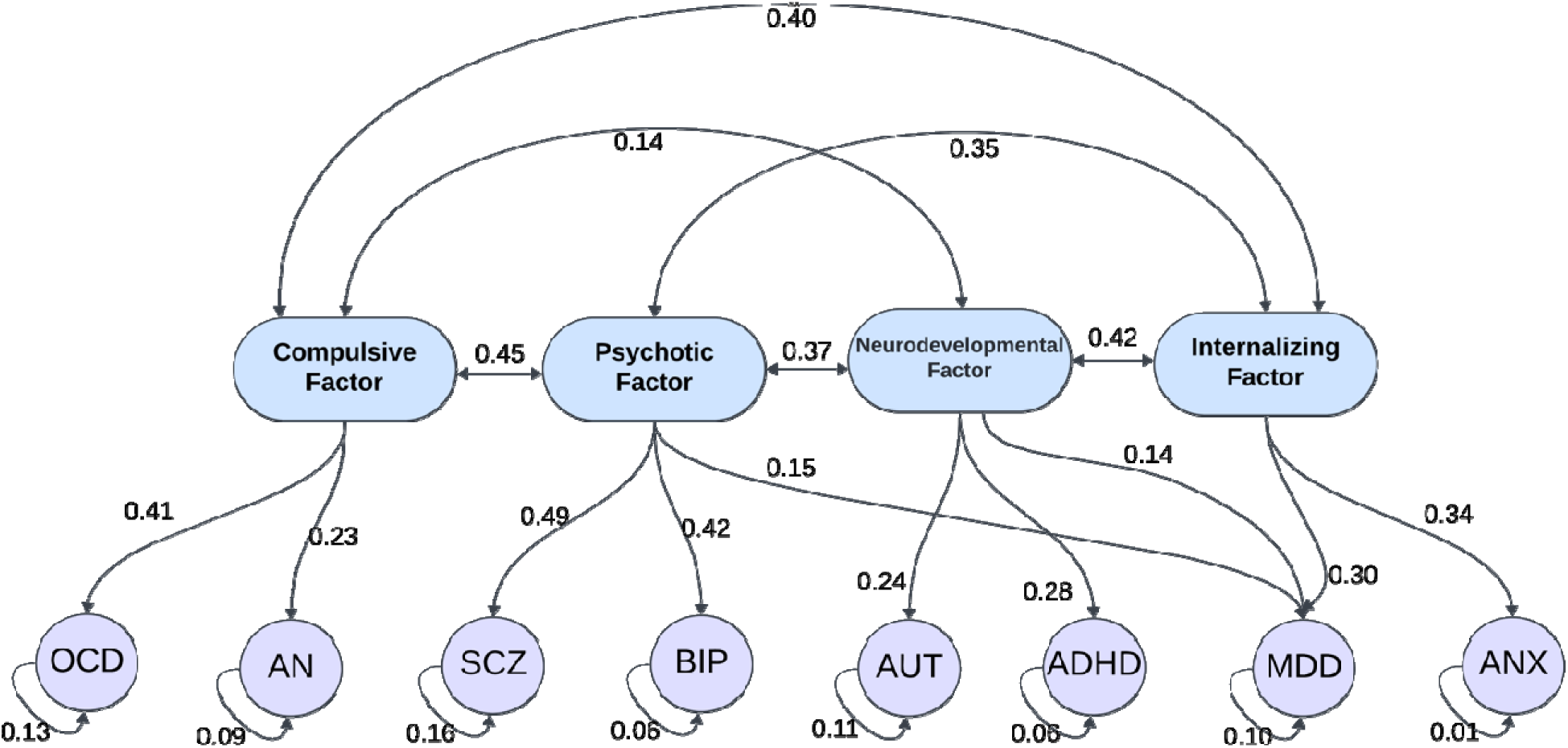
Structural equation model illustrating the hierarchical relationships among latent psychopathology factors and Polygenic risk scores (PRSs) of psychiatric disorders. Four higher-order latent factors—Compulsive, Psychotic, Neurodevelopmental, and Internalizing—are modeled with directional paths between them, reflecting their interrelations. Each latent factor loads onto a subset of *PRSs of* psychiatric disorders: Obsessive-Compulsive Disorder (OCD) and Anorexia Nervosa (AN) load onto the Compulsive Factor; Schizophrenia (SCZ) and Bipolar Disorder (BIP) onto the Psychotic Factor; Autism Spectrum Disorder (AUT) and Attention-Deficit/Hyperactivity Disorder (ADHD) onto the Neurodevelopmental Factor; and Major Depressive Disorder (MDD) and Anxiety Disorders (ANX) onto the Internalizing Factor. Curved double-headed arrows represent correlations or covariances between latent variables and residuals.

After deriving four latent factors based on eight psychiatric disorders, we performed latent factor GWAS on the summary statistics to derive the coefficient weights for factor specific PRSs. GenomicSEM was used to estimate the mean effect sizes of SNPs across disorders, given the latent factor model we specified, resulting in the beta weights for a given SNPs for each of the latent factor. We then deployed those weights to the genetic data of ABCD, generating four PRS for each participant, i.e. COM PRS, PSY PRS, ND PRS, and INT PRS.

### Statistical Analysis

Generalized linear mixed-effects models (GLMMs) were fitted with negative binomial link functions using the R package NBZIMM (Zhang, Yi, & Zhou, 2022). The raw scores of our outcomes (CBCL and BPM-Y) ranged from 0 to 64 and were zero-inflated; distributions of the scores are presented in Appendix Figure S2, S3 and S4; clinical cutoffs are based on t-scores. Therefore, we used a negative binomial link function to model these as dependent variables in mixed-effect regressions. Repeated measures over time and the sibling relatedness of the cohort were modeled using random effects of participant and family, while the independent variables of interest and potential confounding variables were included as fix effects. The independent variables of interest were: 1) indicators for pandemic states (early or late), the reference group is the pre-pandemic state, and 2) the four genetic liability scores (COM PRS, PSY PRS, ND PRS, and INT PRS). The corresponding interactions between pandemic state and genetic liabilities were also included as fixed effects. Other covariates controlled in the fixed effect are baseline collection date, sex-at-birth, age at assessment, household income level, parents’ marriage status, and first ten genetic principal components (PCs) derived from GWAS data with the Random effect to be Family ID nested within ID. Age at assessment was included to control the natural developmental trend of CBCL and BPM-Y scores, as youth gets older. All PRSs and age are scaled and centered. Further stratified analysis on male and female were conducted on the European-like sample.

To improve the interpretability of the results given the negative binomial mixed-effects models, we report the association of the independent variable on the dependent variable via the rate ratio (RR). The RR quantifies the fold change in the rates of the outcome, given one-unit increase in the predictor. The corresponding 95% confidence intervals were calculated for RR and the significant thresholds for hypothesis testing are set at the 0.05 level after Bonferroni correction of 20 tests.

## Results

### Trajectories of mental health during the COVID-19 pandemic

Given wide range of age entering pandemic among ABCD participants (11∼ 14 years in March 2020, Appendix Figure S5), we can model the effects of pandemic period as independent of age-specific effects. The full models were summarized in **Figure 2**. During the early pandemic, a broad range of youth psychopathologies significantly increased, with CBCL and BPM-Y scores rising by approximately 8% to 14% while controlling for polygenic risk scores (PRSs) and age (Figure 2). In the late pandemic, symptom levels increased further—ranging from 8% to 24% above early-pandemic levels—suggesting a potential cumulative effect of prolonged pandemic-related stressors. The most evident late-pandemic increases were CBCL withdrawn/depressive symptoms (18% increase, 95% CI 11% -25%, p =1.299 ×10^-7^), CBCL rule-breaking behavior (21% increase, 95% CI 14% -29%, p =8.545 ×10^-11^), and self-reported BMP-Y internalizing problems (24% increase, 95% CI 19% -30%, p =3.172 ×10^-24^).

**Figure 2:**
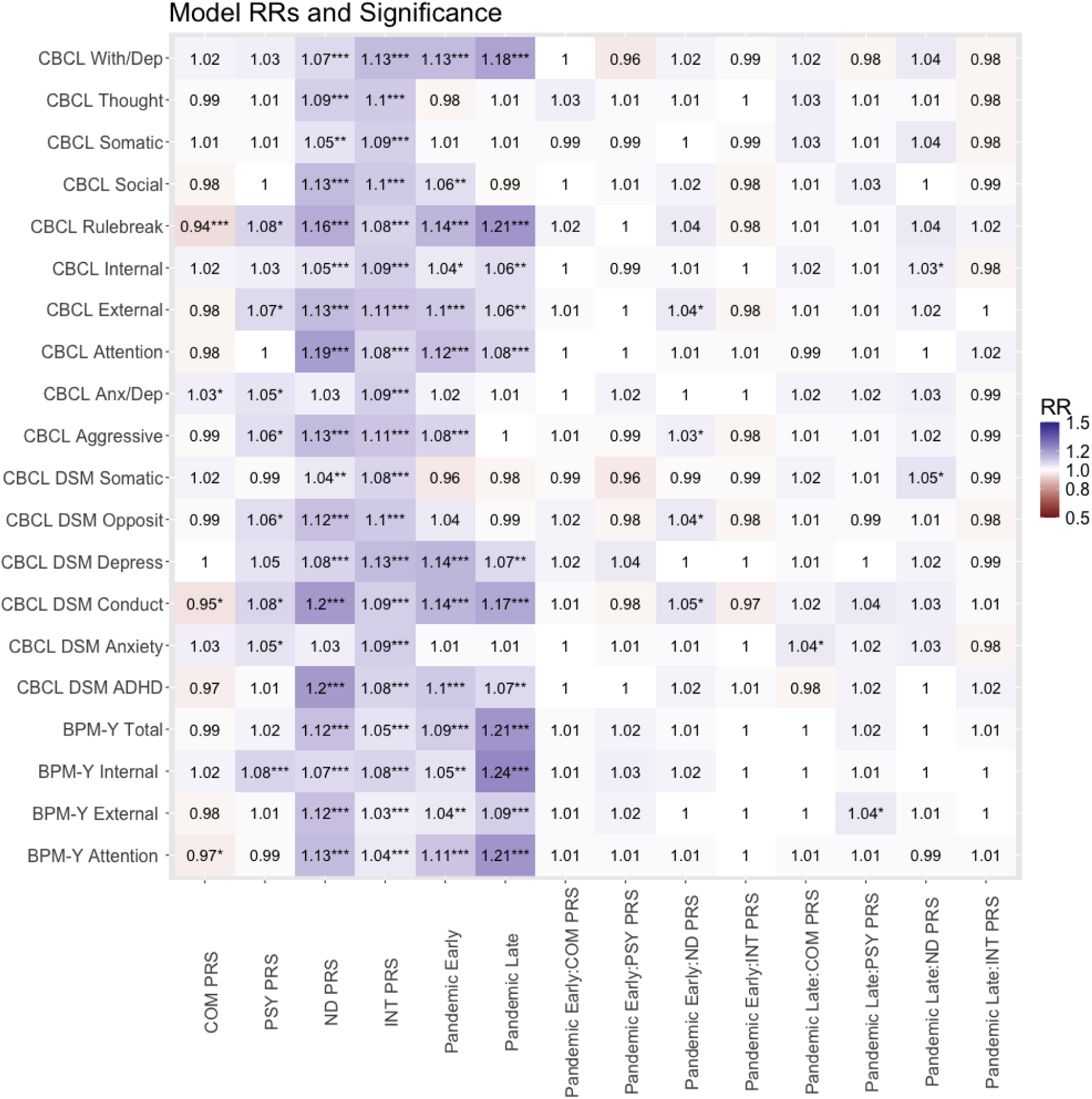
Associations between psychopathologies and the pandemic state and PRSs for European-like set. RR, rate ratio. P-value significance levels: α = 0.05, * indicates that P-value < α, ** indicates FDR corrected significance, *** indicates Bonferroni corrected significance. Each row represents a model. A total of 20 models controlling for baseline collection date, sex-at-birth, age at assessment, household income level, parents’ marriage status, and first ten genetic principal components (PCs) derived from GWAS data with the Random effect to be Family ID nested within ID. Reference group for pandemic states is pre-pandemic period.

### The contribution of pre-disposing vulnerabilities among youth

We tested if our latent factor PRSs were significantly associated with mental health among youth and if the pandemic related worsening of psychopathologies were further accelerated among those who were vulnerable. **Figure 2** summarized the models including all four latent factor PRS and the pair-wise interaction terms, in EUR subset, after controlling for age, family relatedness, and repeated measures. The COM PRS was negatively associated with CBCL rule-breaking problems (6% decrease, 95% CI 2% to 10% decrease, P=1.509×10^-3^). The PSY PRS was significantly associated with BPM-Y internalizing problems (8% increase, 95% CI 4% to 13%, P=5.484×10^-5^). The ND PRS was significantly associated with most of the psychopathologies except for the two-anxiety measures, with 5 to 19 percent more symptoms for every one standard deviation increases of the ND PRS. The INT PRS was significantly associated with all psychopathologies with 3% to 16% increment.

No significant interactions were found between PRS factors and either pandemic period, after Bonferroni correction (all interaction terms: rate ratios ranging from 0.98 to 1.04, P > 0.05). Limited evidence for the multiplicative interactions suggests that pandemic effects on youth’s mental health were compounded uniformly across groups, regardless of their pre-existing vulnerabilities.

### Sex differences in mental health trajectories

We examined the difference in psychopathological changes during the pandemic between male and female participants by sex-stratified analyses. **Figure 3** is the summary of the results, separated into domains of psychopathologies. Across psychopathologies, pandemics have stronger associations with the increasing rates of psychopathologies among females, as evident in the rightward shift of the effects in **Figure 3** (also see Appendix Figure S5, Table S1). Depressive symptoms (CBCL DSM Depress) were higher (**Figure 3a**) among female participants during the early pandemic state (21% increase, 95%CI 13% to 29%, P=2.6×10^-8^), compared to males’ non-significant effect size (1.06, 95% CI 0.995 to 1.12, P=0.07421). The observed sex differences were even more prominent in late pandemic period. During the late pandemic, female participants had higher increment of CBCL Withdraw Depression (25% increase, 95% CI 16% to 35%, P=8.387×10^-9^) and CBCL Rule-breaking (30% increase, 95% CI 19% to 41%, P=2.16×10^-9^). The association of the late pandemic and self-reported BPM-Y scores of female participants also generally exhibited larger effect size than in the analogous male’s models. In contrast, the increase in symptoms among males during early pandemic period was diminished during the late pandemic state. Some mental health issues among males were even reported significantly less by parents than pre-pandemic period (Figure 3).

**Figure 3:**
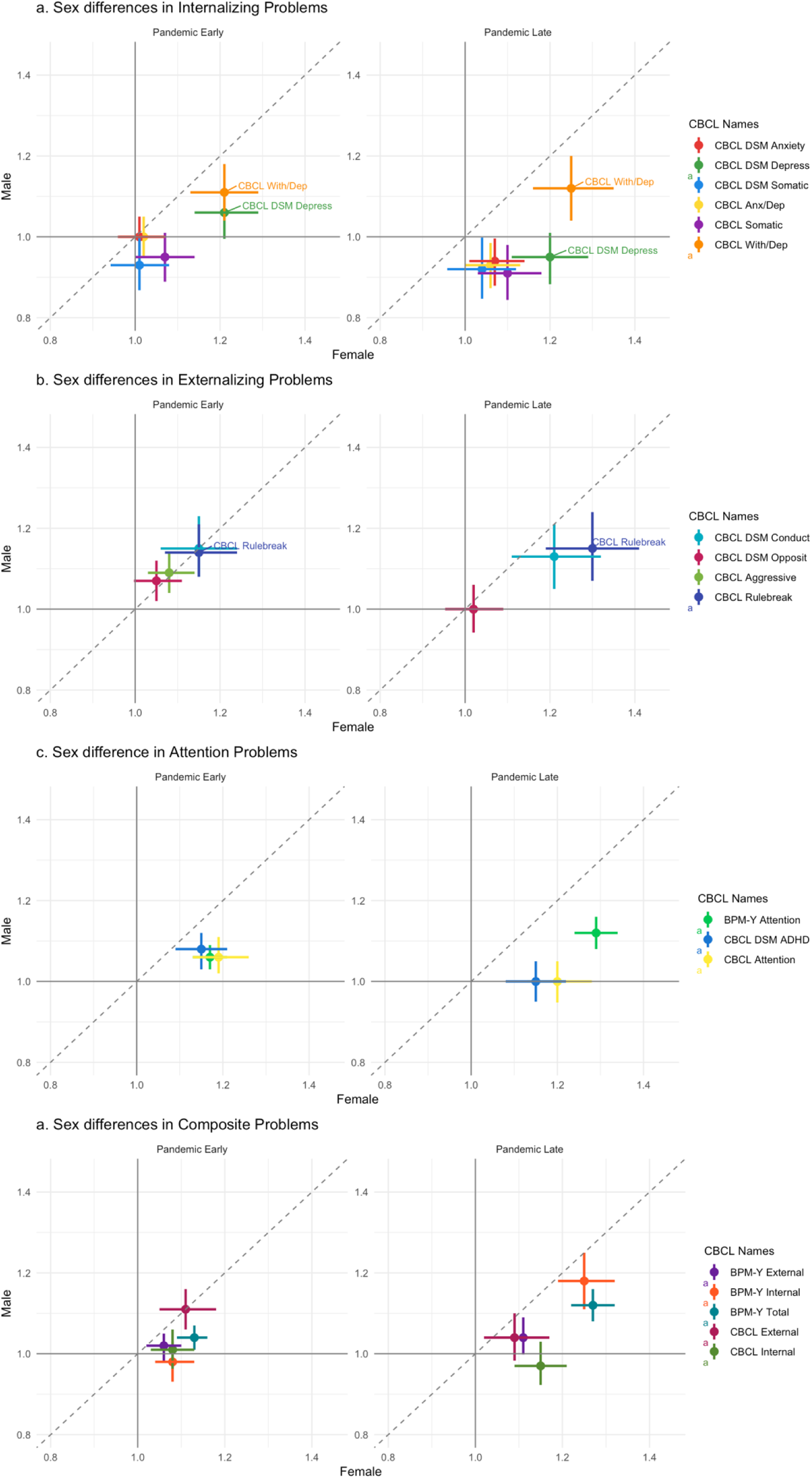
Sex differences in psychopathology across pandemic periods (main effects). Dots represent effect sizes from female models (x-axis) and male models (y-axis), with 95% confidence intervals shown as error bars. The reference group for pandemic state comparisons is the pre-pandemic period. The y-axis displays male-specific effect sizes (rate ratios), and vertical lines denote their confidence intervals. The diagonal dashed line indicates parity between male and female estimates (i.e., Male = Female). Points above the diagonal suggest higher scores in males, while points below suggest higher scores in females. The figure includes: (a) Internalizing problems, (b) Externalizing problems, (c) Attention problems, and (d) Composite problems.

While most of the models show no evident interactions between PRS and pandemic periods, there are also notable differences between males and females. The CBCL DSM depress score was more strongly associated with INT PRS among females (15% increase, 95% CI 10% to 21%, P=6.4×10^-10^) than among males (10% increase, 95% CI 6% to 15%, P=4.2×10^-6^). Meanwhile, with every one standard deviation increase in ND PRS among females, the increasing vulnerability in ND would exhibited more conduct problems during the pandemic periods (**Figure S2, the interaction terms**) (Conduct problem, Early Pandemic*ND PRS, 15% increase, 95% CI 7% to 23%, P=5.166×10^-5^; Aggressive behavior, Early Pandemic *ND PRS, 9% increase, 95% CI 4% to 14%, P=5.085×10^-4^).

### Complexity in diverse population

Among DIV subset, instead of broad range of reported increases in psychopathologies among EUR participants, the early-pandemic-period-related increases in symptoms were limited to the CBCL Withdrawn Depression (11% increase, 95% CI 3% to 20%, P=8.844×10^-3^), CBCL DSM Depression scores (17% increase, 95% CI 8% to 26%, P=8.822×10^-5^), BPM-Y internal (11% increase, 95% CI 6% to 17%, P=4.952×10^-5^) problems, and attention problems (10% increase, 95% CI 6% to 14%, P=3.649×10^-7^). The trending upward maintained during late pandemic period. However, as expected due to higher uncertainty in PRS, fewer PRS associations were found among DIV subset. Only ND PRS is consistently associated with psychopathologies, comparing to those found in the EUR subset (**Figure 4**).

**Figure 4:**
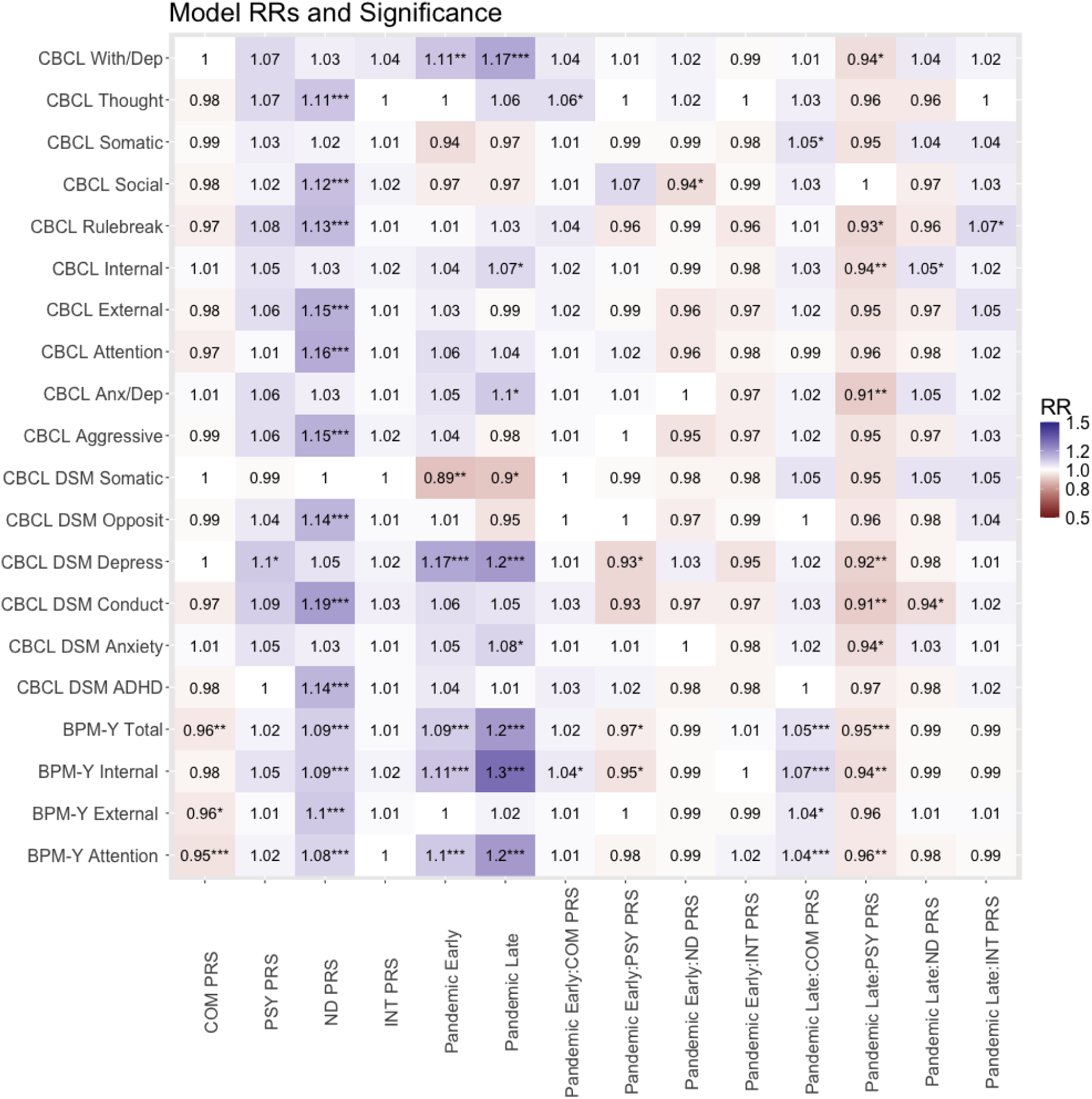
Associations between psychopathologies and the pandemic state and PRSs for diversity(non-European-like) set. RR, rate ratio. P-value significance levels: α = 0.05, * indicates that P-value < α, ** indicates FDR corrected significance, *** indicates Bonferroni corrected significance. Each row represents a model. A total of 20 models controlling for baseline collection date, sex-at-birth, age at assessment, household income level, parents’ marriage status, and first ten genetic principal components (PCs) derived from GWAS data with the Random effect to be Family ID nested within ID. Reference group for pandemic states is pre-pandemic period.

## Discussion

In this study, we found that the associations of the pandemic with the mental wellbeing of youth was persistent and lingering. In adolescent females, the associations were found stronger and longer lasting than in adolescent males. Genetic liabilities for neurodevelopmental and internalizing disorders were associated with increased levels of psychopathology among youth. Although no significant interaction was found, the non-linearity of the negative binomial model implies compounding effects of pandemic exposure and genetic risk by default.

Longitudinal studies have consistently demonstrated that mental health problems originating in childhood can persist and manifest as more severe disorders in adulthood (Solmi et al., 2022). In our analysis, we also found that age at collection time had significant positive associations with almost all psychopathologies. Notably, participants’ ages ranged from 11 to 14 years old in the year 2020. Adjusting for age at each time point accounts for developmental changes in mental health symptoms for everyone without introducing confounding, as age does not influence the likelihood of pandemic exposure. Because psychopathology levels often change with age—especially in youth—this adjustment ensures that observed differences during pandemic are not simply due to participants getting older. In addition, controlling for the baseline evaluation date and site location accounts for cohort differences.

This designed feature allowed us to highlight a progressive intensification of mental health problems among adolescents during the COVID-19 pandemic, with evidence suggesting that symptom exaggeration increased over time. While early-pandemic increases were substantial, the continued rise in symptoms during the late-pandemic period—particularly in withdrawn/depressive symptoms, rule-breaking behavior, and internalizing problems—points to a potential cumulative burden of prolonged stress and disruption.

Importantly, our findings reveal that individual differences in genetic vulnerability moderated these symptom trajectories. Higher polygenic risk for neurodevelopmental (ND) and internalizing (INT) disorders was broadly associated with elevated symptom levels, suggesting that youth with these genetic predispositions may be especially sensitive to environmental stressors such as the pandemic. Notably, the PSY PRS was linked to greater internalizing problems, while the COM PRS was unexpectedly associated with lower rule-breaking behavior, raising questions about potential resilience or compensatory mechanisms within certain risk profiles.

In our negative binomial GLMM, the interaction between the pandemic and polygenic risk scores (PRSs) was not statistically significant. However, the positive main effects of neurodevelopmental (ND) and internalizing (INT) PRSs indicate that genetic vulnerability was associated with greater mental health symptom burden. The effect measure of pandemic period is rate ratio; therefore, it means those with higher baseline symptoms would have fold increment in their symptoms during pandemic. Because the pandemic is associated with accelerating of symptom levels—disproportionately affecting those with higher baseline symptoms—youth with elevated genetic risk experienced greater absolute increases. This suggests that, even without a statistical interaction, genetic vulnerability compounded the mental health concerns during the COVID-19 pandemic. An example is given in Figure 4; just considering the main effects, the increments in BPM-Y Internalizing problem scores during the pandemic for those with 2 standard deviation higher ND PRS is 70% higher for those with 2 standard deviation lower ND PRS. These findings highlight the importance of considering both genetic and environmental factors to better identify and support at-risk youth.

**Figure 4:**
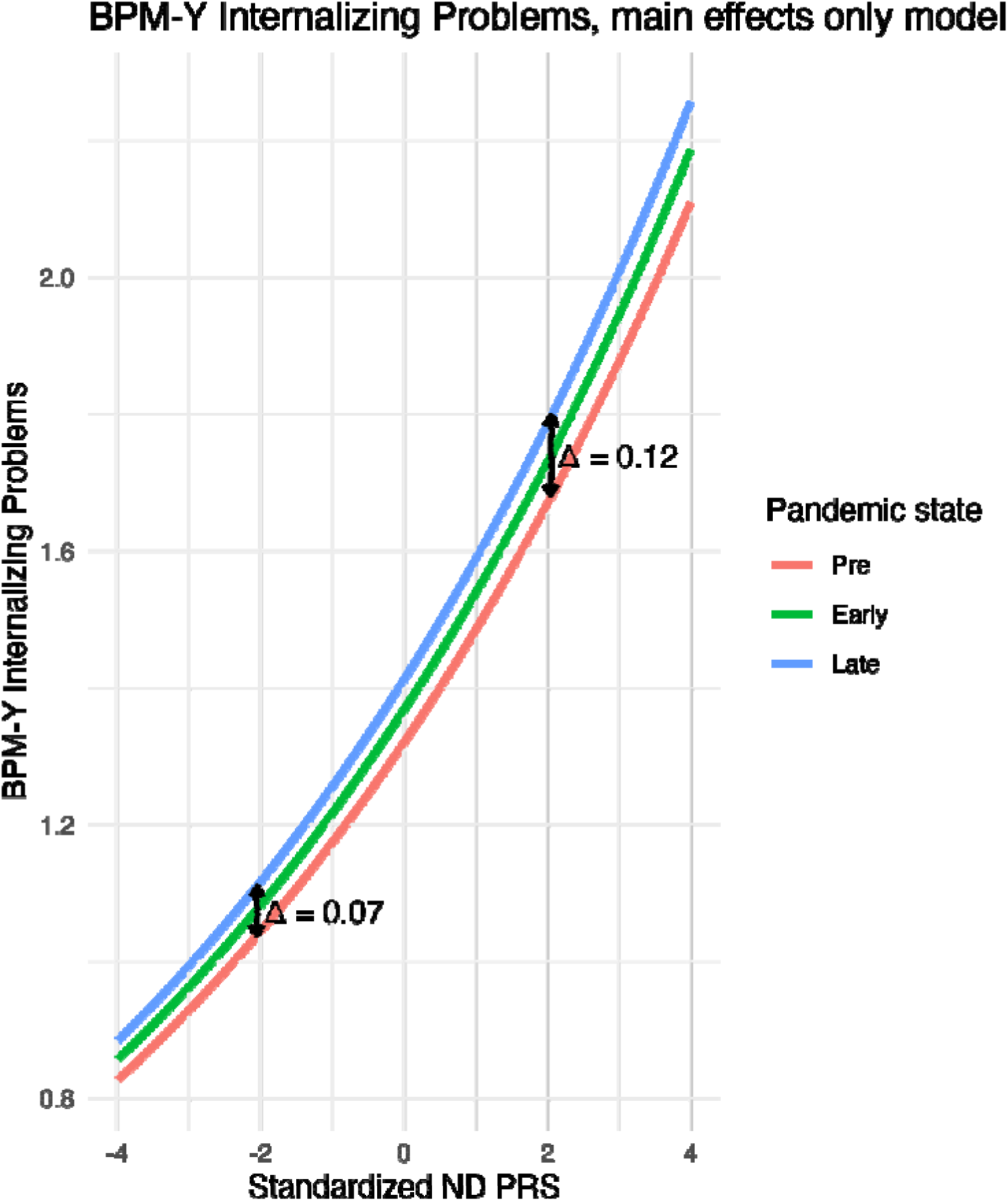
Visualization of main effects only without the interaction term of ND PRS and the pandemic on BPM-Y Internalizing problems. Model fitted BPM-Y Internalizing problems across levels of standardized neurodevelopmental polygenic risk scores (ND PRS), stratified by pandemic status. The figure shows main effects only, with other covariates held at their mean (for continuous variables) or reference level (for categorical variables). Subjects with higher standardized ND PRS show a proportionally greater increase in BPM Internalizing problems during the pandemic, despite the absence of an explicit interaction term in the model.

Loades et al. (2020) found that the absence of social connections during adolescence can hinder emotional development and exacerbate mental health vulnerabilities. Their review suggests that loneliness is linked to increased depression in girls and social anxiety in boys, with girls being particularly sensitive to perceived social exclusion—an experience shown to have lasting effects on well-being (Prinstein & Giletta, 2016). Our findings align with this literature: during the COVID-19 pandemic, adolescent females experienced a sustained decline in mental health, likely driven by prolonged social isolation. In contrast, mental health concerns among adolescent males appeared to stabilize during the later phases of the pandemic. These findings underscore the need for sex-specific approaches to mental health prevention and care. Additionally, our study shows that individuals with genetic risk for neurodevelopmental disorders remain especially vulnerable to varies mental problems, even outside of pandemic conditions. This highlights the importance of early intervention and sustained, individualized support for at-risk youth. More broadly, these insights call for a shift in how public health policies address mental health—especially during crises. Beyond medical or genetic vulnerabilities, social support, community connection, and equitable access to mental health resources must be prioritized. Future public health strategies should integrate these social dimensions to better protect the mental well-being of vulnerable populations, particularly in times of widespread disruption.

There were limitations in our study. The lack of specific measurements on COVID-19 related social disruptions makes it difficult to fully understand the various ways in which the pandemic affected adolescents’ mental health. Generalized data often miss the complexity of disruptions such as school closures, changes in peer interactions, and the shift to virtual communication, which may have impacted youth differently. Without more targeted measures that account for these factors, it becomes challenging to assess the direct relationship between social disruption and mental health outcomes, hindering effective interventions. Limited data for individuals of non-European-like ancestry introduces uncertainty in the findings for these groups, as reduced sample sizes limit statistical power and may affect the generalizability and reliability of genetic associations. This underrepresentation underscores the need for more inclusive genomic research to ensure equitable understanding and application of genetic risk across diverse populations. While the CBCL (parent-report) and BPM-Y (self-report) offer complementary perspectives on youth mental health, each has limitations—CBCL may miss internal experiences, while BPM-Y can be influenced by self-report biases. Differences in informant perspective and symptom visibility may account for variations in findings. However, the use of both measures across multiple time points strengthens the longitudinal analysis by capturing diverse and evolving patterns of psychopathology from multiple viewpoints.

## Conclusion

Adolescent mental health worsened over the course of the COVID-19 pandemic, showing cumulative increases in symptom burden, and this study uniquely examined these influences using comprehensive measures, including psychopathology as assessed by the CBCL and BPM-Y, while rigorously controlling for multiple genetic factors via PRSs and developmental changes via age. This approach allowed us to adjust for inherent genetic vulnerabilities and examine potential genetic interactions with pandemic-related stress, potentially offering deeper insights into the interplay between genes and environment. By exploring potential gene-by-environment interactions, we found that although both genetic vulnerabilities and pandemic-related challenges contributed to increased symptoms, no direct interaction was observed between the two in general. However, because rate ratios are multiplicative, the presence of two significant main effects implies a naturally proportional increase in symptoms, even in the absence of an interaction. Pronounced sex differences showed female adolescents experienced more severe mental health declines than males. Individuals with neurodevelopmental genetic vulnerabilities emerged as particularly at risk for severe mental health declines.

## Data Availability

All data produced in the present study are available upon reasonable request to the authors

## Supporting Information

Table S1: Models of psychopathologies and the pandemic state and PRSs stratified by sex.

## Acknowledgements

This work was supported by grant R01MH122688, RF1MH120025, and R01128959, funded by the National Institute for Mental Health (NIMH). Data used in the preparation of this article were obtained from the Adolescent Brain Cognitive Development SM(ABCD) Study (https://abcdstudy.org), held in the NIMH Data Archive (NDA). The ABCD Study® is supported by the National Institutes of Health and additional federal partners under award numbers U01DA041048, U01DA050989, U01DA051016, U01DA041022, U01DA051018, U01DA051037, U01DA050987, U01DA041174, U01DA041106, U01DA041117, U01DA041028, U01DA041134, U01DA050988, U01DA051039, U01DA041156, U01DA041025, U01DA041120, U01DA051038, U01DA041148, U01DA041093, U01DA041089, U24DA041123, U24DA041147. A full list of supporters is available at https://abcdstudy.org/federal-partners.html. A listing of participating sites and a complete listing of the study investigators can be found at https://abcdstudy.org/consortium_members/. ABCD consortium investigators designed and implemented the study and/or provided data but did not necessarily participate in the analysis or writing of this report. This manuscript reflects the views of the authors and may not reflect the opinions or views of the NIH or ABCD consortium investigators. The ABCD data repository grows and changes over time.

## Key points

- This study investigates how patterns of exaggerated reporting across a broad range of mental health problems intensified over time during the COVID-19 pandemic, while accounting for individual genetic risk for various classes of mental disorders in youth.
- Both genetic vulnerabilities are associated with increased psychiatric symptoms. Individuals with neurodevelopmental and internalizing genetic vulnerabilities (assessed using PRSs) were particularly at risk of severe mental health problems.
- By exploring potential gene-by-environment interactions, we found that although both genetic vulnerabilities and pandemic-related challenges contributed to increased symptoms, no direct interaction was observed between the two in general.
- This analysis also revealed pronounced sex differences, with female adolescents experiencing more severe mental health declines than their male counterparts during the pandemic.

## Appendix

**Figure S1:**
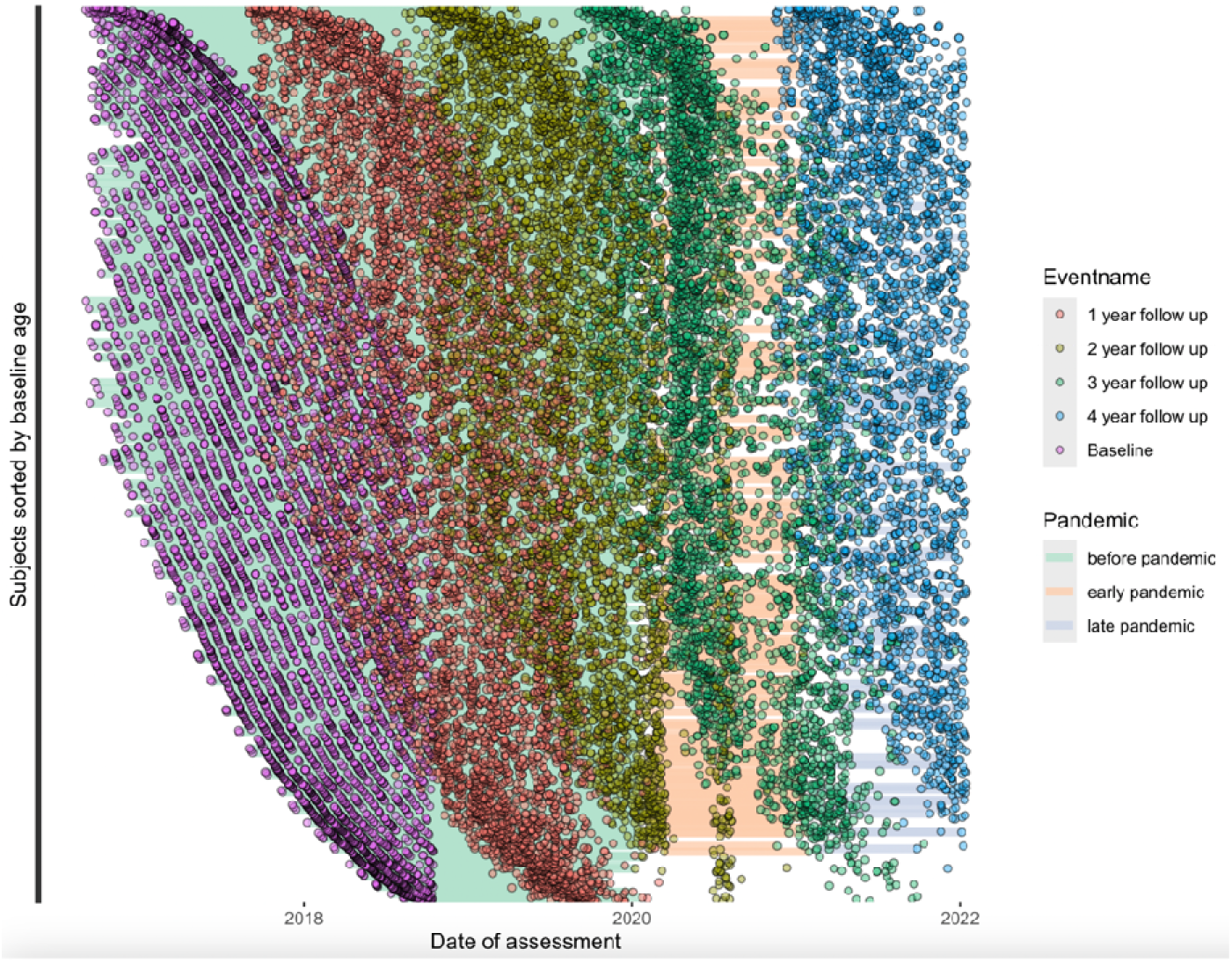
Outcomes collection time visualization sorted by baseline age, with five events.

**Figure S2:**
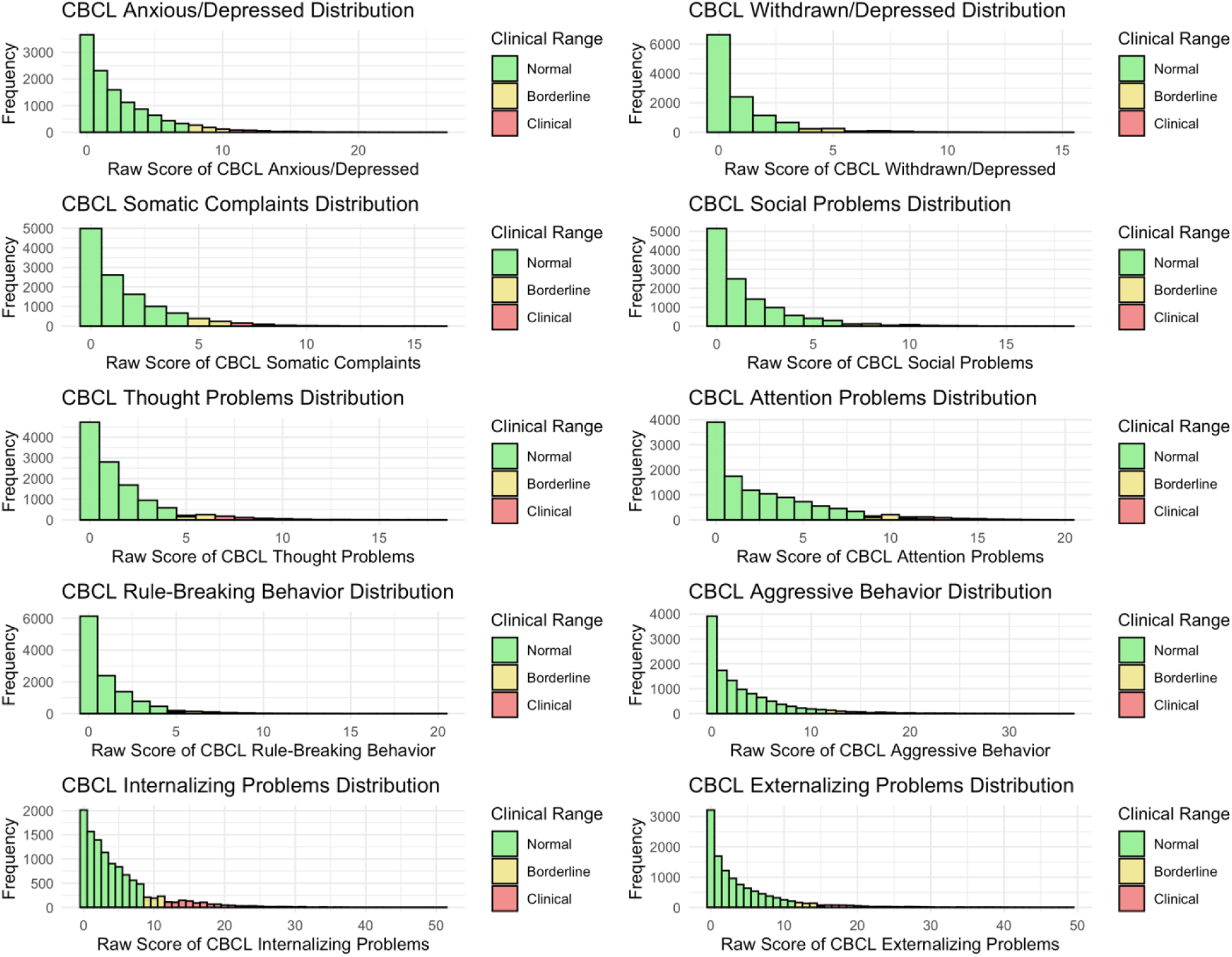
Distribution of CBCL Syndrome scores (raw scores). Borderline and Clinical ranges are based on the corresponding t-scores.

**Figure S3:**
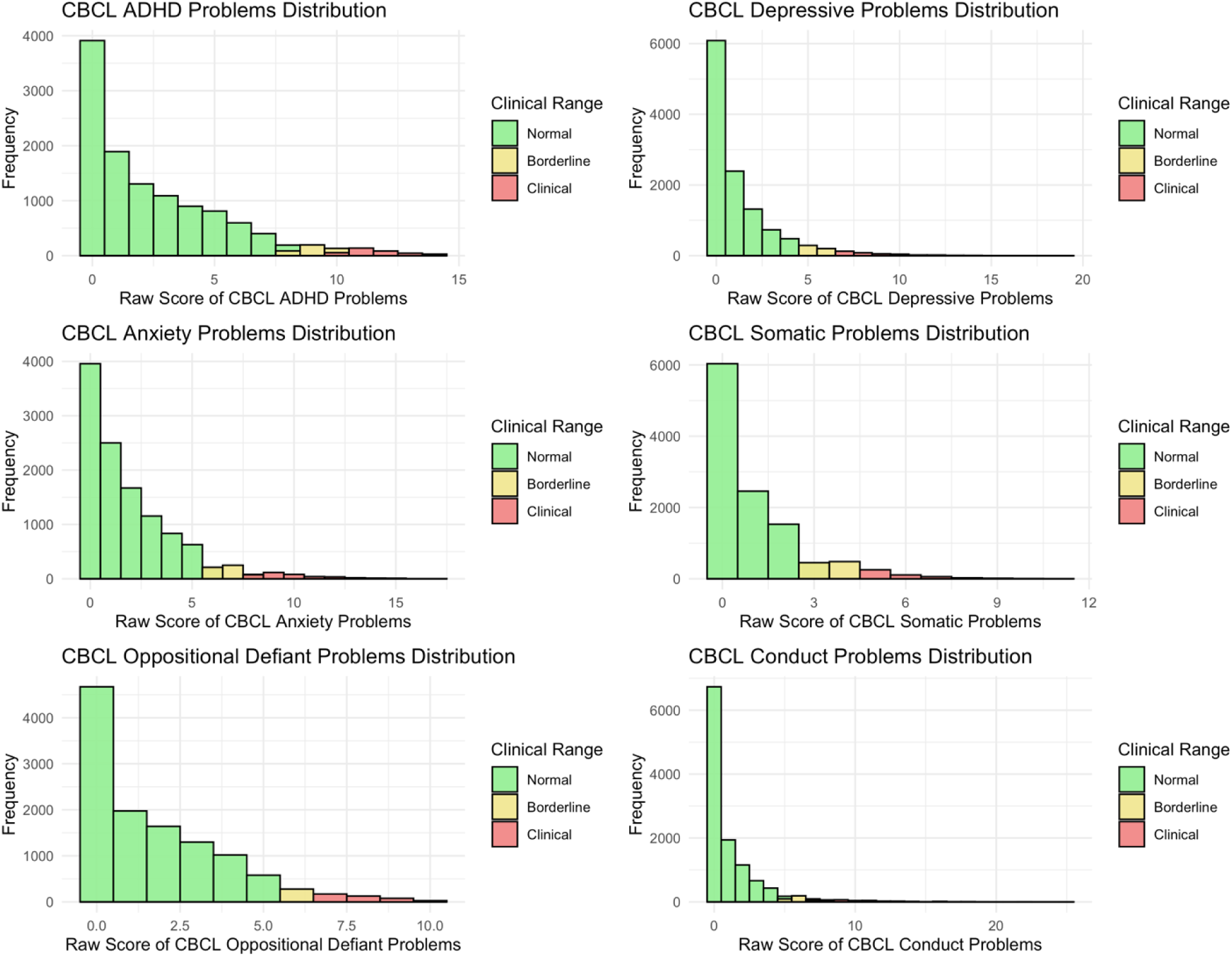
Distribution of CBCL DSM-oriented scores (raw scores). Borderline and Clinical ranges are based on the corresponding t-scores.

**Figure S4:**
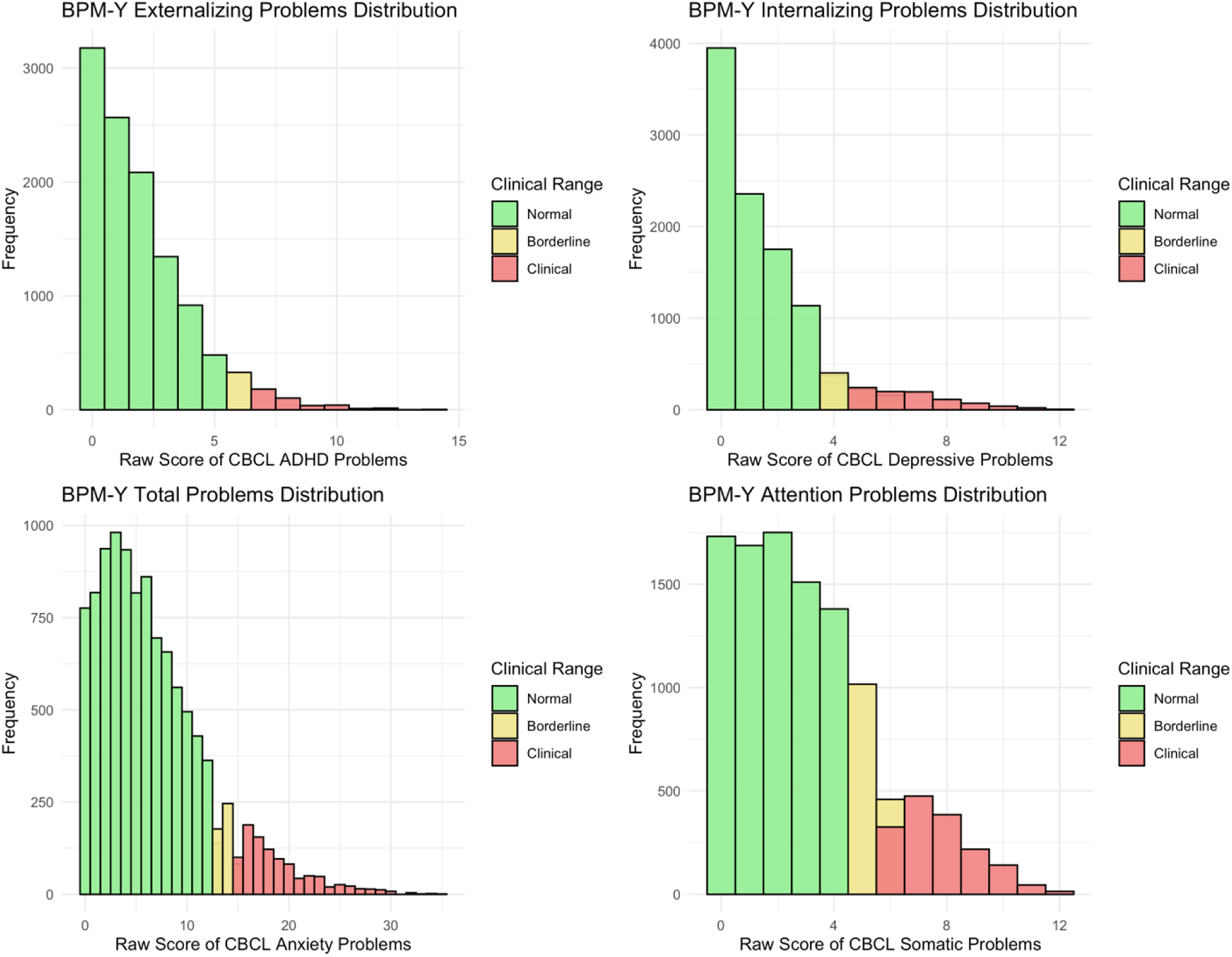
Distribution of BPM-Y scores. Borderline and Clinical ranges are based on the corresponding t-scores.

**Figure S5:**
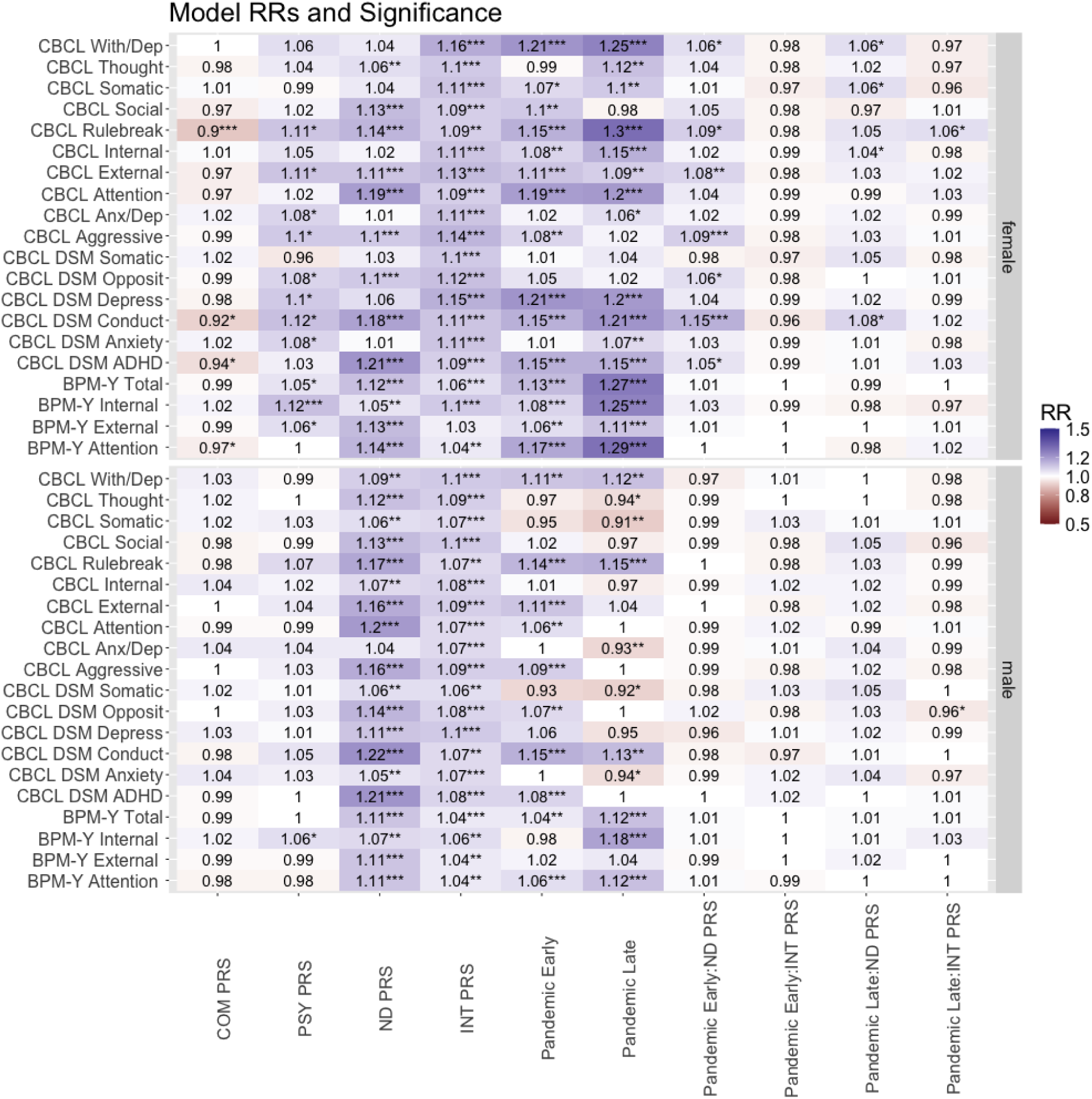
Associations between psychopathologies and the pandemic state and PRSs stratified by sex. RR, rate ratio. P-value significance levels: α = 0.05, * indicates that P-value < α, ** indicates FDR corrected significance, *** indicates Bonferroni corrected significance. Each row represents a model. A total of 40 models adjusted for sex, age at interview, site location, and first 10 principal components (PCs) derived from GWAS data with the Random effect to be Family ID nested within ID.

